# Reanalysis of cluster randomised trial data to account for exposure misclassification using a per-protocol and complier-restricted approach

**DOI:** 10.1101/2023.04.20.23288835

**Authors:** Suzanne M. Dufault, Stephanie K. Tanamas, Citra Indriani, Riris Andono Ahmad, Adi Utarini, Nicholas P. Jewell, Cameron P. Simmons, Katherine L. Anders

## Abstract

The intention-to-treat (ITT) analysis of the Applying *Wolbachia* to Eliminate Dengue (AWED) trial estimated a protective efficacy of 77.1% for participants resident in areas randomised to receive releases of *w*Mel-infected *Aedes aegypti* mosquitoes, an emerging dengue preventive intervention. The limiting assumptions of ITT analyses in cluster randomised trials and the mobility of mosquitoes and humans across cluster boundaries indicate the primary analysis is likely to underestimate the full public health benefit. Using spatiotemporally-resolved data on the distribution of *Wolbachia* mosquitoes and on the mobility of AWED participants (n = 6,306), we perform complier-restricted and per-protocol re-examinations of the efficacy of the *Wolbachia* intervention. Increased intervention efficacy was estimated in all analyses by the refined exposure measures. The complier-restricted analysis returned an estimated efficacy of 80.7% (95% CI: 65.9, 89.0) and the per-protocol analysis estimated 82.7% (71.7, 88.4) efficacy when comparing participants with an estimated *w*Mel exposure of *≥* 80% compared to those with *<*20%. These reanalyses demonstrate how human and mosquito movement can lead to underestimation of intervention effects in trials of vector interventions and indicate that the protective efficacy of *Wolbachia* is even higher than reported in the primary trial results.

## Introduction

A breakthrough in efforts to curtail the global spread of dengue has recently come in the form of the intracellular bacterium *Wolbachia* (*w*Mel strain), which increases the resistance of *Aedes aegypti* mosquitoes – the primary vector of the dengue virus – to the replication and onward transmission of a number of arboviral diseases including dengue, Zika, chikungunya, and yellow fever^1–4^. Encouraging results from quasi-experimental field trials indicated that introgression of *w*Mel into local *Ae. aedes* populations was associated with reduced incidence of notified dengue cases^5–7^. The efficacy of *Wolbachia* for dengue control was demonstrated experimentally in a gold standard parallel-arm cluster randomised trial (CRT) in Indonesia (the AWED trial, ‘Applying *Wolbachia* to Eliminate Dengue’), which reported a protective efficacy of 77.1% (95% CI: 65.3%, 84.9%) against virologically-confirmed dengue in the primary intention-to-treat (ITT) analysis.^8^

While highly promising, the ITT estimate is unlikely to capture the full intervention effect due to aspects of the study design that are not well handled by an ITT analysis, resulting in an underestimated intervention efficacy^9^. In cluster randomised trials of community-delivered interventions, such as the *w*Mel deployments in the AWED trial, individual participants’ true exposure status may be different to the cluster-allocated intervention status due to mobility of humans and spillover of the intervention across cluster boundaries. *w*Mel coverage is heterogeneous across time and space, within “intervention” clusters themselves and with observed spillover into “untreated” clusters during the later portion of the study period. Further, humans move outside of their cluster of residence in their daily routines. When such mosquito and human mobility extends the protective effect of the intervention to individuals in the control clusters, and dilutes the exposure of ostensibly ‘treated’ individuals who spend time in control clusters, it can result in traditional ITT analyses that underestimate the full public health benefit.

A first effort at moving beyond the ITT to examine the full protective effect of the *Wolbachia* intervention was a pre-specified secondary ‘per-protocol’ analysis of the AWED trial data in which participants’ exposure status was reclassified from a cluster-level binary to an individual-level weighted ‘*Wolbachia* Exposure Index’ (WEI) to account for self-reported mobility and measured differences in *w*Mel establishment across the study area. In this initial ‘per-protocol’ analysis, reported together with the ITT analysis in the publication of trial results^8^, the WEI was estimated in two distinct ways: 1) based solely on the measured cluster-level *w*Mel prevalence in *Ae. aegypti* mosquitoes in the participant’s cluster of residence during the calendar month of participant enrolment, and 2) based on the cluster-level *w*Mel prevalence in each cluster the participant reported visiting in the week prior to illness onset, weighted by the proportion of total observed time the participant spent in each cluster. The methods are further described in the AWED study protocol^10^. The WEI based on cluster of residence alone resulted in very little exposure variability (i.e. most participants had WEI values near 1 or 0), making it difficult to perform reliable risk comparisons at intermediate levels of exposure. Dengue risk was significantly reduced in only the highest WEI stratum (WEI*≥*80%) compared to the lowest (WEI *<* 20%), with efficacy estimated at 77.4% (95% CI: 62.1%, 85.6%), very similar to the ITT efficacy estimate. The WEI based on *w*Mel and travel history had a bit more variation in exposure for participants in each arm, and estimated a dose-response effect, suggesting statistically significant differences in protective efficacy for individuals with at least a WEI of 40% as compared to those with WEI less than 20%^8^.

Both definitions of WEI used a somewhat crude estimation of time spent under protective *w*Mel cover, relying on cluster-level *w*Mel proportions in the month of enrolment; a more geographically and temporally localised *w*Mel proportion may more accurately capture an individual’s protective exposure. Simulation work by others demonstrates that a greater protective efficacy is expected than observed in the initial ITT and per-protocol analyses^9^. Human and mosquito mobility occur on much finer scales than the 1 km^2^ geographic regions (“clusters”) randomised to treatment allocation. Ignoring the contamination effect of this movement has been shown to bias the efficacy estimate towards the null^9^.

The present study re-estimates AWED trial participants’ individual-level *w*Mel exposure using spatially and temporally resolved data on the distribution of *Wolbachia* mosquitoes and on trial participants’ mobility collected during the AWED trial, in order to perform an improved per-protocol and a complier-restricted re-examination of *Wolbachia* efficacy in the AWED trial. This granular exposure data accounts explicitly for human mobility, as well as the spatial heterogeneity in *w*Mel coverage particularly arising from spillover of *w*Mel mosquitoes across the boundaries between treated and untreated clusters. By reducing misclassification in individuals’ *Wolbachia* exposure status, these reanalyses of data from the AWED cluster randomised trial provide the most unbiased experimental estimate to date of the true efficacy of *Wolbachia* for dengue control.

## Results

### Human mobility

*w*Mel-infected *Ae. aegypti* mosquitoes were released into 12 intervention clusters (of 24 total clusters) in Yogyakarta between March and December 2017. Patients presenting to local health clinics (*puskesmas*) between January 2018 and March 2020 with undifferentiated acute febrile illness of 1 to 4 days duration and aged between 3 and 45 years were invited to enrol in the AWED trial^10^. Those who consented to enrol had a blood sample collected for dengue diagnostic testing and were asked about their movements during daytime hours (5am – 9pm) in each of the 3 to 10 days prior to their illness onset, corresponding to the incubation period of a dengue virus infection. The 6306 participants included in the AWED analysis dataset reported spending the majority of their time at home (Fig. 1, median 68.8%, interquartile range 57.8-85.2%), consistent with the results of a baseline mobility survey in Yogyakarta^11^.

**Figure 1.**
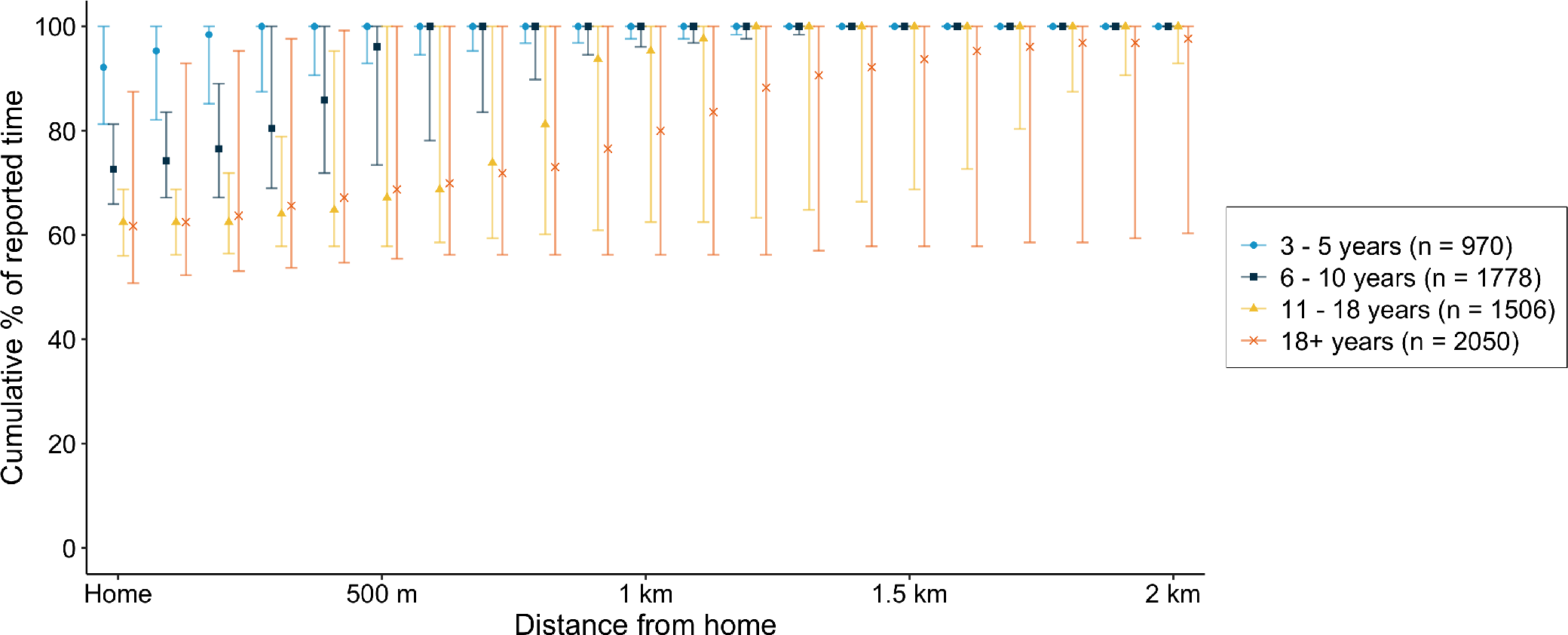
Mobility of AWED trial participants in Yogyakarta City during the 3-10 days prior to illness onset, based onself-reported travel (5AM - 9PM). The graph shows the median and interquartile range of the time participants within each age group spent at home and cumulatively at increasing distances from home (children aged 3-5 years, 6-10 years, 11-18 years, and greater than 18 years).

Though 87.2% of participants left their residence at least once during the 8-day period (Fig S1A), typically to attend school (56.8%) or work (19.2%) (Fig S2), 86% of the reported locations visited were within 1km of a participant’s residence (Fig S3). The number of unique locations visited did not differ between adults and children, between study arm of residence, or between dengue cases and test-negative controls (Fig S1B-D). However, the aggregate time that participants spent at increasing distances from home increased with age, with only 28.5% (276/970) of young children 3-5 years and 32.1% (571/1778) children 6-10 years reporting any time spent further than 1km from home, compared to 54.5% (821/1506) adolescents 11-18 years and 64.9% (1330/2050) adults. More than a third of participants (39%) stayed within their cluster of residence throughout all 8 days. A median of 98.4% (IQR 68.0-100.0%) of participants’ time was spent under the same intervention arm as their cluster of residence. The proportion of time under the intervention assignment showed no apparent differences for individuals in the intervention versus untreated arm.

### *w*Mel heterogeneity

Adult mosquitoes were collected from a fixed network of 455 traps approximately monthly throughout the 27-month trial period in order to monitor *w*Mel prevalence in *Ae. aegypti* over time in the intervention clusters and contamination in untreated clusters. Between January 2018 and March 2020, there were a total of 10,432 mosquito trapping events, only 10 of which had no *Ae. aegypti* present. A total of 87,679 *Ae. aegypti* mosquitoes were screened; 49,266 (56.19%) of which were detected to have *w*Mel.

The majority of BG traps were located within the AWED RCT study boundaries (Fig. 2A), with 181 traps in the intervention region and 186 traps in the untreated region. An additional 11 traps were located in an untreated area on the southeast boundary of the RCT study site and 77 traps in a *Wolbachia*-treated area on the northwest boundary of the RCT study site; these areas had served as the untreated control and intervention area, respectively, in a quasi-experimental *Wolbachia* release study prior to the AWED randomised trial^5^. Overall, 96.1% and 93.6% of mosquitoes screened in the quasi-intervention and RCT intervention areas contained *w*Mel, respectively. Only 9% of the mosquitoes captured in the quasi-untreated and 15% of mosquitoes from the RCT untreated areas had *w*Mel present. In the RCT intervention and quasi-intervention areas, the marginal monthly capture and detection of *w*Mel in mosquitoes was fairly consistent across time. In the RCT untreated area, there was a marked increase in the proportion and absolute number of *w*Mel-infected mosquitoes detected in the final year of the study (Fig. 2B).

**Figure 2.**
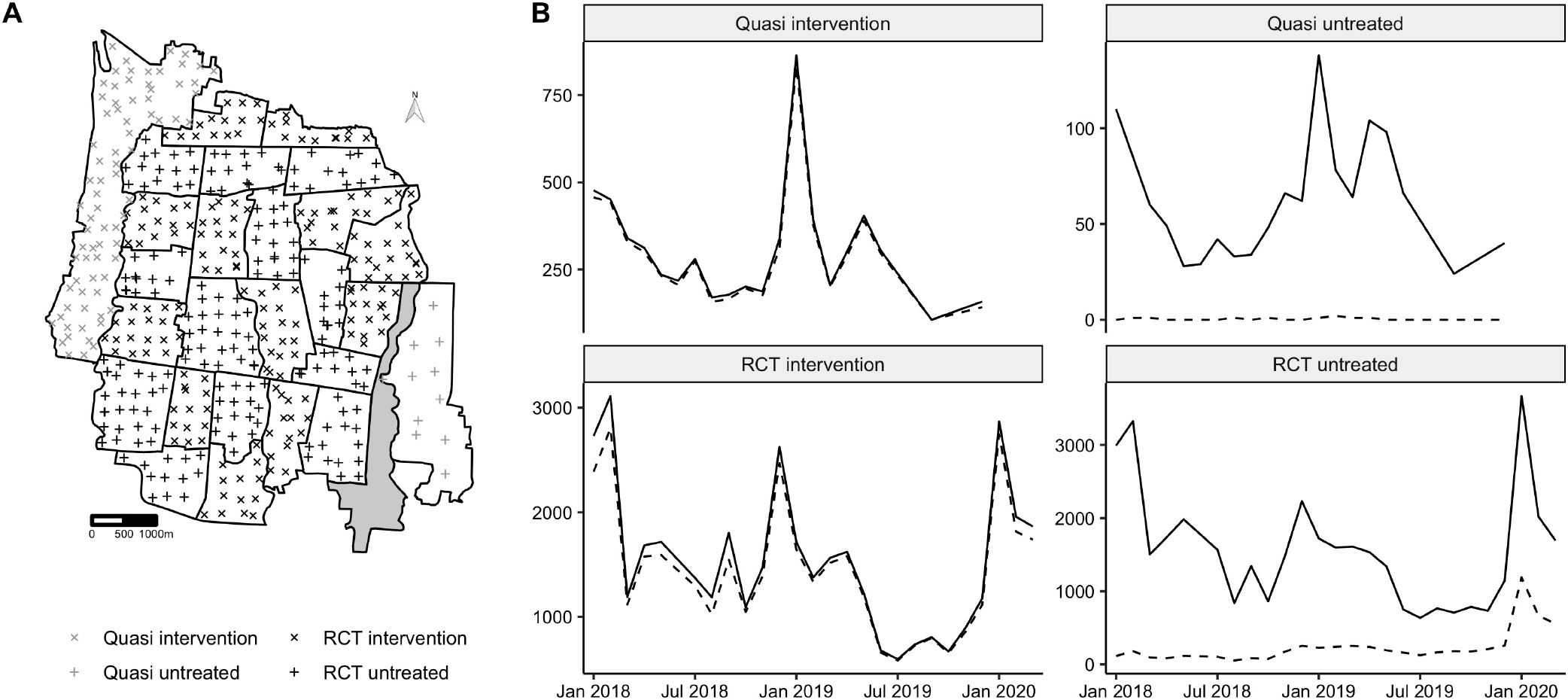
(A) Map of Yogyakarta City with geolocated trap locations. Shape is used to distinguish intervention areas where *w*Mel releases occurred (‘X’) and untreated areas where *w*Mel releases did not occur (‘+’). Traps within the AWED RCT study boundaries are denoted in black, while those in the quasi-experimental areas are marked in gray. (B) The total number of mosquitoes screened (solid line) and the number of mosquitoes with *w*Mel detected (dashed line) per month by study area.

### Complier-restricted reanalysis of *Wolbachia* efficacy

To examine the efficacy of *Wolbachia* among those who received the intervention as randomly assigned, we restricted the analysis to those participants who reported spending all of the potential DENV exposure period under the intervention assignment concordant with their cluster of residence (“compliers”). Among the 6,306 participants in the AWED primary analysis data set, 3,114 participants (49%) stayed strictly within the RCT study area and within clusters in the same study arm as their cluster of residence, during the 3 to 10 days prior to illness onset: 1,359 test-negatives and 33 dengue cases in the intervention arm and 1,530 test-negatives and 192 dengue cases in the untreated arm. Applying the same modified odds ratio approach used in the primary analysis of the AWED trial^8^ and described in an earlier methods paper^12^, intervention efficacy in the complier-restricted subgroup was estimated at 80.6% (95% confidence interval: 65.9%, 89.0%), about 3 percentage points higher than that estimated in the primary analysis.

If we expand the definition of compliers to include participants who left the RCT study area but remained in areas with concordant treatment assignment to their cluster of residence, an additional 772 participants are included: 64 additional test-negatives in the intervention arm and 651 test-negatives and 57 dengue cases in the untreated arm. Intervention efficacy in this subgroup was similarly estimated at 79.7% (95% confidence interval: 67.2%, 87.4%).

### Per-protocol reanalysis of *Wolbachia* efficacy

To examine the protective effect of *Wolbachia* exposure on dengue risk, regardless of compliance with intervention assignment, we constructed an interpolated *w*Mel surface to estimate each individual participant’s *Wolbachia* exposure level. Figure 3 shows the inverse density weighted maps of *w*Mel coverage aggregated monthly across a 100 *×*100 m grid overlaid on the AWED study site. Across each panel, it is evident that *w*Mel is established at a high level (light regions) in intervention areas and, as suggested by Figure 2, begins to creep across the boundaries between the intervention and untreated regions by years 2 and 3 of the trial, leaving the centers of the untreated clusters uncontaminated (dark regions). The results from the leave-one-out cross-validation method used to determine the optimal hyperparameters for interpolation are provided in the Supplemental Material.

**Figure 3.**
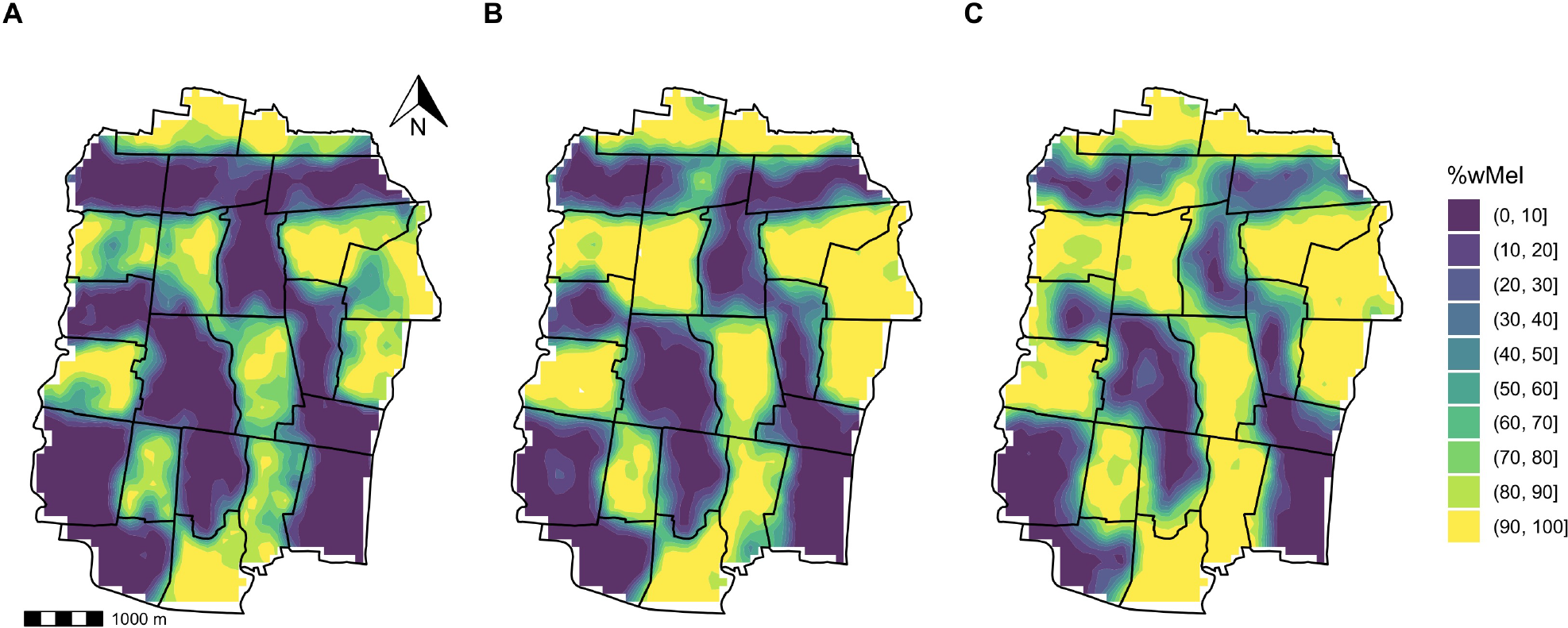
Spatiotemporal inverse density weighted maps of *w*Mel% interpolated across a 100x100 meter grid overlaid on the AWED trial surface on January 1 each year of (A) 2018, (B) 2019, and (C) 2020.

We approach per-protocol re-estimation of efficacy in two ways, mirroring the previous efforts towards obtaining individual-level WEI^8,10^. We first estimate an individual’s *Wolbachia*-exposure index (WEI) based on the spatiotemporally interpolated proportion of *w*Mel mosquitoes at the individual’s residence on the reported day of illness onset, referred to as ‘interpolated WEI (residence)’. Second, we use a weighted sum of the individual’s interpolated WEI over their travel history and residence in the 3 to 10 days prior to illness onset, referred to as ‘interpolated WEI (activity)’. A moving sum of the ultimate and penultimate trap collections (relative to a participant’s illness onset) was used in both interpolation procedures in order to stabilise the highly variable individual trap collections.

The per-protocol reanalysis based on the interpolated WEI (residence) estimated the *Wolbachia* efficacy to reach 82.0% (95% CI: 72.3%, 87.9%) when comparing those whose residential WEI was above 0.8 to those whose residential WEI was below 0.2 (Figure 4). Figure 4 further shows that a monotonic dose-response relationship with efficacy was observed as the interpolated WEI (residence) increased. Statistical significance was found at every contrasting WEI (residence) strata when WEI was at or above 0.4. A similar trend was observed with interpolated WEI (activity), where statistically significant effects were observed in every WEI stratum, with a maximum efficacy estimate of 82.7% (95%CI: 71.7%, 88.4%) for WEI greater than 0.8 and minimum efficacy estimate of 33.7% (95% CI: 21.7%, 46.5%) observed at WEI within [0.2, 0.4), compared to WEI less than 0.2 (Figure 4). Compared to the original per-protocol results based on cluster-aggregate WEI, the reanalysis using interpolated values of individual WEI resulted in an increase in the estimated intervention efficacy and (for the WEI based on residence only) a more pronounced dose-response relationship.

**Figure 4.**
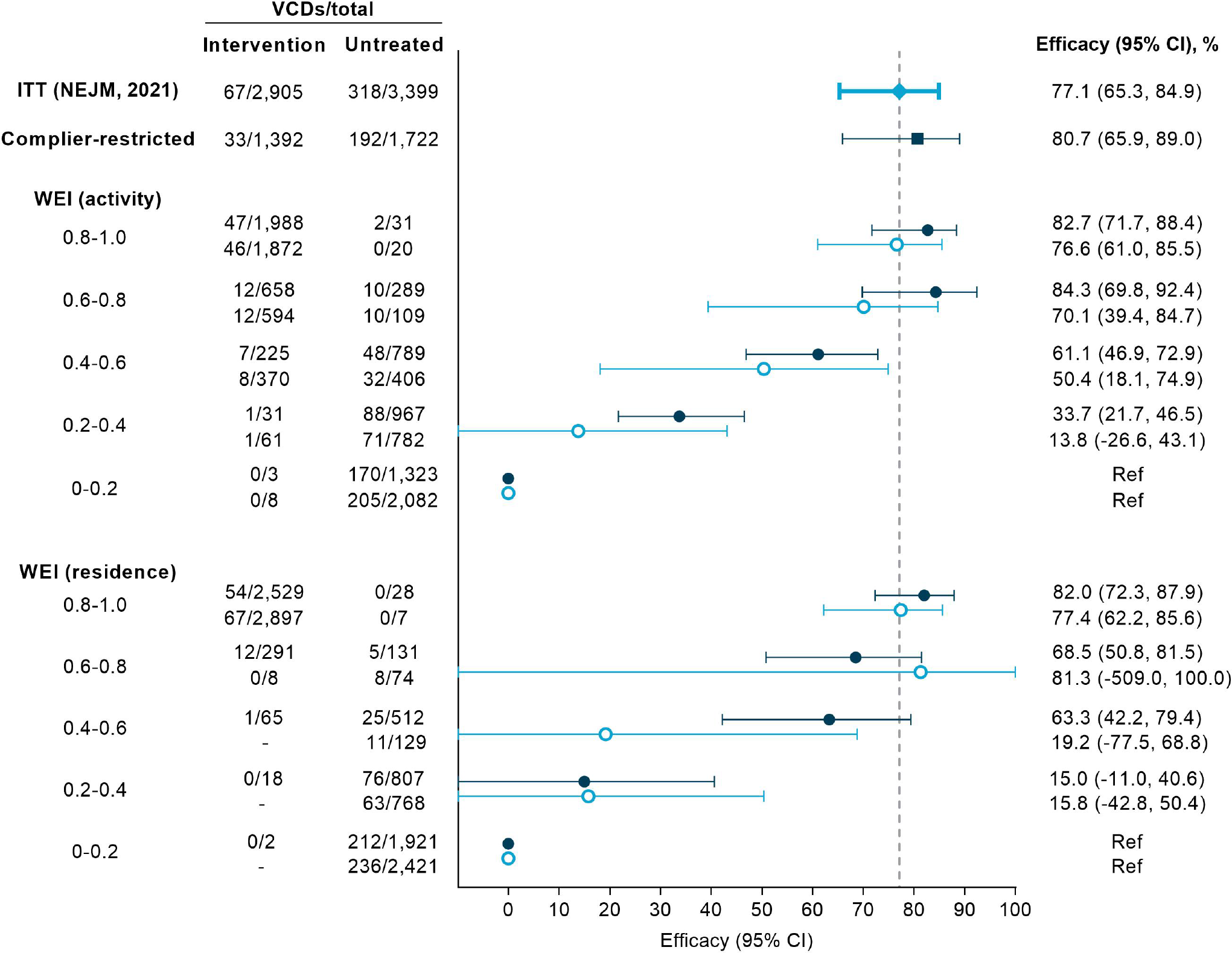
Forest plot comparing *Wolbachia* efficacy estimates from the primary analysis and reanalysis of the AWED trial in Yogyakarta, Indonesia. Included are the original ITT estimates^8^ (light blue diamond), the complier-restricted analysis results performed here (dark blue square), the new per-protocol estimates based on spatio-temporally interpolated WEI (navy, closed circle), and the original per-protocol estimates based on the cluster-aggregate WEI (light blue, open circle).

## Discussion

After accounting for human movement and intervention contamination across cluster boundaries, our reanalysis of data from a cluster randomised trial of *Wolbachia*-infected mosquito releases for the control of dengue showed an increased estimate of intervention efficacy compared to the primary analyses published previously. Participants’ observed movement patterns were strongly age-dependent, highlighting the need for consideration of human mobility in both the design and – where possible– the analysis of trials for interventions delivered at a community level. Substantial spillover of *Wolbachia* mosquitoes into several untreated clusters during the 27-month period of clinical enrolment complicated the measurement of the intervention effect, but also suggests that small initial gaps in *Wolbachia* coverage under programmatic deployment could fill themselves in with time without further intervention.

In randomised controlled trials, misclassification of participants’ exposure can occur if they do not receive – or are non-compliant with – the intervention they were randomly allocated to receive. This exposure misclassification produces an estimate of intervention efficacy that is biased towards the null when the comparison groups are more similar in their exposure status than dictated by the randomised treatment allocation. In the specific case of cluster-randomised trials of area-level interventions without buffer areas between clusters, contamination by human movement and/or spillover of the intervention across cluster boundaries can lead to an underestimate of efficacy in the ITT analysis. In the AWED trial, most participants reported spending the majority of their time prior to illness onset under their intervention assignment, however half of participants spent at least 15% of their time either outside the trial area or in a location with a discordant intervention assignment. In addition, a marked increase in *w*Mel-infected mosquito spillover into untreated clusters was observed in the final year of the trial. These contamination risks were recognised at the design stage in the AWED trial, in which 12 of 24 contiguous clusters were randomised to receive releases of *Wolbachia*-infected mosquito releases for control of dengue^10^, but could not be accounted for fully in the initial intention-to-treat or per-protocol analyses, which relied on cluster-level summary values of *Wolbachia* prevalence in *Ae. aegypti* mosquitoes. By deriving spatiotemporally interpolated values of *Wolbachia* prevalence at participants’ homes and other visited locations, and combining these with detailed travel history data to calculate individual-level WEI in order to reduce biases in exposure measurement, we have estimated the intervention efficacy to be more than five percentage points higher than in the ITT analysis (82.7% [95%CI 71.7, 88.4] vs 77.1% [65.3, 84.9]), among participants with a *Wolbachia* exposure index *>*80% .

The results of our reanalysis suggest that exposure misclassification from both human mobility and spillover of *w*Mel mosquitoes contributed to the underestimate of *Wolbachia* intervention effect in the ITT analysis. In the complier-restricted analysis, the point estimate of intervention effect increased by three percentage points compared to the ITT analysis after removing only the biasing effect of human mobility by restricting the analysis to those participants who spent all of their time within the assigned intervention arm, while still considering cluster-level *Wolbachia* status as a binary (treated vs untreated). The further increase in efficacy measured in the per-protocol reanalysis – both relative to the ITT analysis and to the original per-protocol analysis based on cluster-level *w*Mel prevalence – indicates that within-cluster variability in *w*Mel prevalence was also contributing to the misclassification of individual participants’ *w*Mel exposure status. The estimated intervention effect for participants in the highest stratum of *w*Mel exposure (80-100%), relative to the lowest stratum (0-20%) was very similar regardless of whether or not an individual’s exposure was based on interpolated *w*Mel prevalence at their primary residence only, or weighted by time spent at other visited locations. This observation further supports the home as the primary location of dengue transmission risk, at least in Yogyakarta city.

Recent work^13^ used geolocated residence, date of illness onset, and DENV serotype information to estimate small-scale spatiotemporal dependence among the dengue cases detected in the AWED trial and showed, for the first time, that the focal clustering of dengue cases is interrupted by the presence of *w*Mel-infected mosquitoes. Those results support the proposition that the true *w*Mel intervention effect was larger than measured in the primary analysis of the AWED trial.

While this work aimed to reduce exposure misclassification by taking a finer spatiotemporal snapshot of an individual’s WEI, lingering complications in estimation persist. Fogelson et al (forthcoming) find that estimates of intervention efficacy from mixed effects models, rather than the generalised lineal models applied here, can be biased when within-cluster exposure levels are highly homogeneous. When this estimator bias is properly handled, their work estimates a greater than 90% intervention effect from the WEI activity and residence individual-level exposures. However, the Fogelson et al analysis modelled WEI as a continuous variable such that the efficacy estimate is based on a contrast of 100% versus 0% WEI, whereas our analysis contrasts the highest (80-100%) versus the lowest (*<*20%) WEI stratum.

Theoretical and empirical studies of *w*Mel invasion into *Ae. aegypti* populations have predicted slow but steady spread at a rate of 100-200 metres per year in northern Australia^14,15^. *w*Mel spread was predicted to be slower in tropical regions with higher *Ae. aegypti* densities, with variation in mosquito population density and geographical barriers contributing to variations in dispersal. This is consistent with the observed spread of *w*Mel across the boundaries of several untreated clusters by 27 months after the end of releases.

Following the successful demonstration in the AWED trial of the efficacy of *w*Mel-infected mosquito deployments in reducing dengue incidence, *w*Mel deployments into the untreated areas of Yogyakarta city were completed between October 2020 and January 2021. Between August 2021 and November 2022, *w*Mel deployments were expanded to the districts of Sleman and Bantul adjacent to Yogyakarta city, reaching a cumulative estimated population of 1.8 million people in a contiguous area of 540 km2. A recent secondary analysis of the AWED trial^16^ has shown that a *Wolbachia* intervention effect equivalent to the AWED result was measurable from interrupted time series analysis of routine dengue case notification data. The area-wide coverage of *Wolbachia* in and around Yogyakarta city now provides an opportunity to evaluate the feasibility for *w*Mel to lead to sustained suppression, or even local elimination (Bannister-Tyrrell et al 2023, in preparation), of dengue in Yogyakarta.

## Methods

### Data source

The Applying *Wolbachia* to Eliminate Dengue (AWED) trial^8, 10^was a cluster randomised, parallel arm study with test-negative sampling carried out in Yogyakarta, Indonesia for a period of 27 consecutive months, beginning in January 2018 and ending during the emergence of the global coronavirus pandemic in March 2020. The city of Yogyakarta was divided into 24 contiguous clusters each approximately 1km^2^, randomised 1:1 to receive releases of *w*Mel-infected mosquitoes between March and December 2017 or no intervention. Routine vector control efforts continued throughout the study in both arms.

### Participant data

Participants enrolled in the AWED trial provided demographic information (age and sex), a geolocated residential address, and a detailed travel history for the hours between 5 AM and 9PM over the three to ten days before illness onset. A total of 116,473 unique movement events were reported by the 6,306 individuals in the analysis sample. The majority of reported locations fell within the RCT study area (92.9%). There were 8,277 movement events outside of the RCT area: 1,091 (*<*1%) and 533 (*<*1%) fell in the quasi-experimental intervention and untreated areas, respectively. The remaining 6,653 (5.7%) events fell outside of any study area.

### Entomological monitoring

A network of 367 BG Sentinel traps (Biogents, Germany) collected adult mosquitoes throughout the AWED trial area between January 2018 and March 2020, such that median trap density was 16.4 BG/km^2^ in the intervention clusters and 15.3 BG/km^2^ in the untreated clusters^8^. An additional 88 BG Sentinel traps monitored *Wolbachia* prevalence in two regions adjacent to the AWED trial site: one comprised of seven kelurahans (urban villages) on the northwestern perimeter of Yogyakarta City, where *Wolbachia*-infected mosquitoes had been released between August 2016 and March 2017 in a quasi-experimental field trial, and a second region comprised of three kelurahans on the southeastern perimeter, which had served as an untreated control area in that trial (Fig. 2A). The study and its results have been described elsewhere^5^. The median trap density was 15.7 BG/km^2^ in the quasi-experimental intervention area and 3.6 BG/km^2^ in the quasi-experimental non-release area (Figure 2A)^5^. All traps were monitored on staggered weekly schedules. The number of mosquitoes caught in each BG trap was recorded by species, sex, and in total. *Ae. aegypti* were stored at -20°C in 80% ethanol until testing for *w*Mel infection^5,8^.

## Descriptive analyses

### Human mobility

Descriptive analyses of participant mobility were performed examining 1) the proportion of cumulative observation time at varying distances from home by age group (Fig. 1), 2) number of unique locations visited (Fig. S1), and 3) type of location (Fig. S2).

### wMel Spatiotemporal Heterogeneity

In the previously reported per-protocol analysis, *w*Mel spatiotemporal heterogeneity was incorporated into the efficacy estimate by taking the proportion of trapped *Ae. aegypti* with *w*Mel *Wolbachia* detected, in either the cluster of residence or as a time-weighted average across the clusters visited in the month of enrolment^10^. Here, we leverage the extensive and frequently monitored BG trap network to obtain estimated *w*Mel proportions at much greater proximity to residential and visited locations.

### Complier-restricted analysis

The complier-restricted analysis restricted the analytic dataset to those who spent all reported time under the intervention assignment determined by their cluster of residence (n = 3,114). Further detail on how this was determined can be found in the Supplemental Material.

Intervention efficacy was estimated with the modified odds ratio approach used in the primary analysis from the NEJM manuscript^8^, as described in the study protocol^10^ and earlier methods paper^12^.

### Per-protocol reanalysis

We used the interpolated *w*Mel surface to estimate individual levels of exposure. Just as in the initial secondary analysis of the AWED trial data^8^, we first estimated an individual’s *Wolbachia*-exposure index (WEI) based on the spatiotemporally interpolated proportion of *w*Mel mosquitoes at the individual’s residence on the reported day of illness onset. Second, we used a weighted sum of the individual’s interpolated WEI over their travel history and residence in the 3 to 10 days prior to illness onset.

Intervention efficacy was estimated with the same approach as in the initial report^8^, using a generalised linear model and balanced bootstrap resampling approach based on cluster residence.

## Data Availability

Participants’ residential address was collected and stored under the ethical approval of the AWED trial protocol. Because the geolocated residence is considered personally identifiable information, individuals seeking access to this data should contact Katherine Anders to discuss obtaining ethical approval or accessing a deidentified version of the dataset. Trap-level entomological data is available for download at the GitHub repository maintained by the first author (https://github.com/sdufault15/yogya-wei-spatial). All analyses were performed with R and the analytical code can be found on GitHub (https://github.com/sdufault15/yogya-wei-spatial).

## Acknowledgements

We acknowledge the contribution of all investigators in the AWED Study Group to the implementation of the trial. We thank the leadership and residents of Yogyakarta for their support and participation in the trial. We gratefully acknowledge the Tahija Foundation as funders of the AWED trial, and the Wellcome Trust and the Bill and Melinda Gates Foundation, which provided financial support to the World Mosquito Program.

## Author contributions statement

All authors were involved in reviewing the final manuscript. S.M.D contributed to the conceptualization, analysis, writing of the original draft, and revision of the final manuscript. S.K.T. contributed to the conceptualization, analysis, writing of the original draft, and revision of the final manuscript. C.I. contributed to the data curation, investigation, and provision of resources. A.U. contributed funding acquisition and project administration. R.A.A contributed resources and project administration. N.P.J. contributed to the conceptualization, analysis, and editing of the manuscript. C.P.S. contributed to the conceptualization, project administration, and editing of the manuscript. K.L.A. contributed to the conceptualization, project administration, writing of the original draft, and revision of the final manuscript.

## Additional information

### Ethics Approval

The trial protocol for the Applying Wolbachia to Eliminate Dengue (AWED) trial study was approved by the Universitas Gadjah Mada (UGM) ethics committee (approval number KE/FK/105/EC/2016) and Monash University Human Research Ethics Committee (approval number 0960). Written informed consent was obtained from participants (or their guardian where the participant is a minor). In addition, participants between 13 and 17 years of age were invited to sign a consent form indicating that they understood the research and agreed to participate.

### Competing interests

We confirm that we have no conflicts of interest to disclose.

## Notes

### Competing Interest Statement

The authors have declared no competing interest.

### Clinical Trial

NCT03055585

### Summary of Updates

This minor revision has an updated abstract and small changes to the main text that should help with clarity around approaches taken and interpretation of results. There have been no major revisions to analyses or data that impact the previously reported results.

