## Supplemental Material for "Reanalysis of cluster randomised trial data to account for exposure misclassification using a per-protocol and complier-restricted approach"

Suzanne M. Dufault, Stephanie K. Tanamas, Citra Indriani, Riris Andono Ahmad, Adi Utarini, Nicholas P. Jewell, Cameron P. Simmons, Katherine L. Anders

#### Contents

**1 Human mobility** **2**

**2 Spatiotemporal Inverse Density Weighting** **3**

**3 Changes in WEI** **4**

**4 Determining the "Compliers" for Complier-Restricted Analysis** **4**

### 1 Human mobility

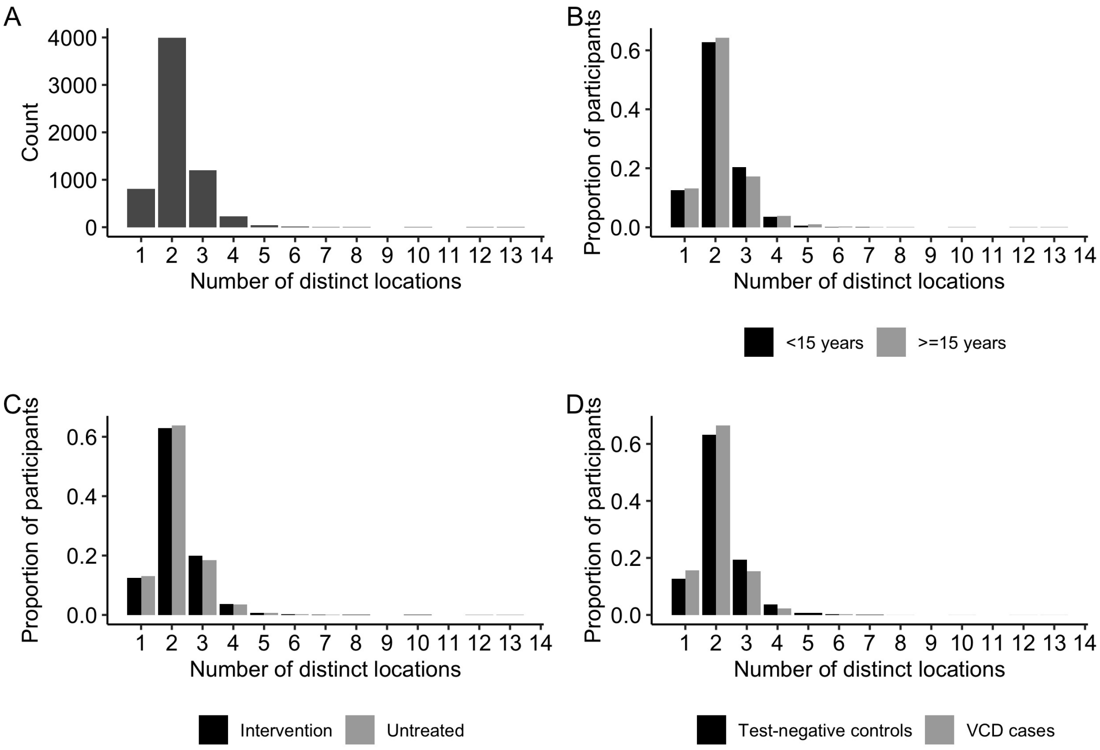


Figure S1: Number of distinct locations AWED trial participants self-reported visiting between 5am and 9pm during the 3-10 days prior to illness onset, including their primary residence. **A)** Frequency distribution of number of visited locations among 6306 participants in the analysis dataset. **B-D)** Relative frequency distribution of number of visited locations among participants $<15$ vs $\geq 15$ years (B), participants resident in intervention vs untreated clusters (C), and virologically-confirmed dengue cases vs test-negative controls (D).

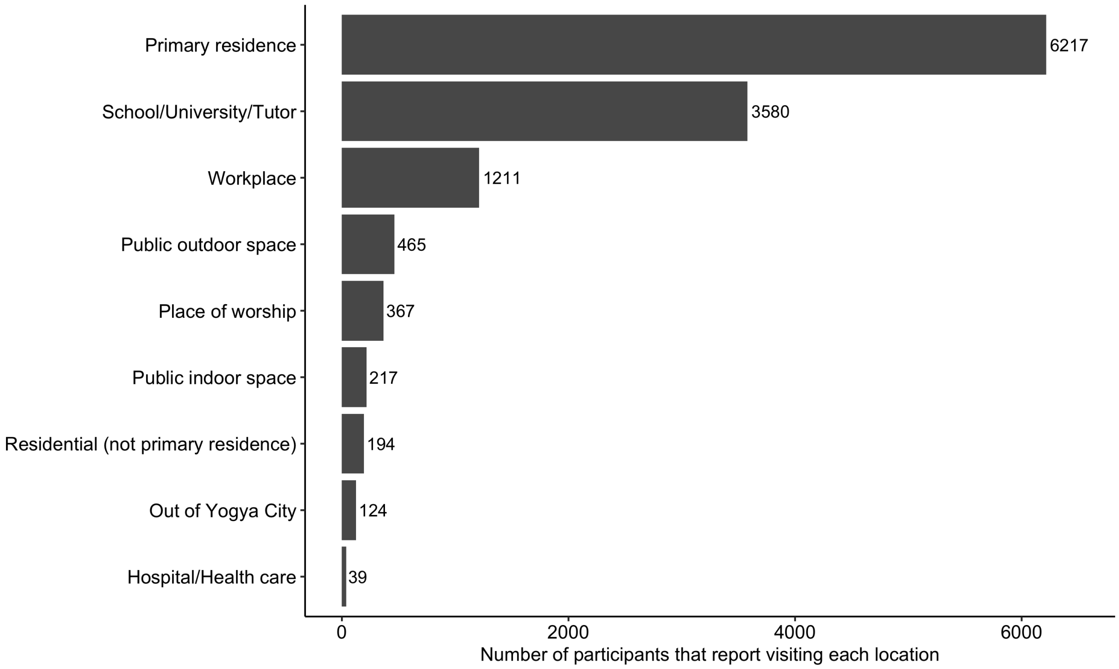


Figure S2: The top locations sorted by the number of participants who reported visiting each location.

### 2 Spatiotemporal Inverse Density Weighting

Before interpolating the $w$Mel surface across the study area, the trap-level proportions of $w$Mel mosquitoes were stabilised by summing the observed mosquito counts and observed mosquito counts with $w$Mel detected for the current and penultimate trap event. Spatiotemporal inverse density weighting (IDW) was used to construct interpolated $w$Mel surfaces.

The general formula for inverse density weighting (IDW) is given in Equation 1, where $\hat{Z}(\cdot)$ is the estimated proportion of mosquitoes with $w$Mel at a specified location $s_0$ and time $t_0$. Let $s_i$ denote the $i$th observed location $\{s_i : i = 1, 2, \ldots, m_T\}$ which has two spatial components $(x_i, y_i)$, for instance, longitude and latitude. Let $t_j$ denote the time of the $j$th observation where observations were made at $\{t_j : j = 1, 2, \ldots, T\}$.

$$\widehat{Z}(s_0; t_0) = \sum_{j=1}^{T} \sum_{i=1}^{m_j} \lambda_{ij} Z_{ij}(s_i; t_j) \tag{1}$$

The multiplier $\lambda_{ij}$ represents the inverse density weight (Eq. 2), which is a function of distance in space-time $(d_{ij,0})$ from the observed data $Z(s_i, t_j)$ and the space-time location of interest $\hat{Z}(s_0, t_0)$.

$$\lambda_{ij} = \frac{d_{ij,0}^{-p}}{\sum_{i=1}^{n} d_{ij,0}^{-p}} \tag{2}$$

Distance in this setting was estimated using Equation 3.

$$d((s_i, t_j), (s_0, t_0)) = \sqrt{(x_i - x_0)^2 + (y_i - y_0)^2 + C \cdot (t_j - t_0)^2} \tag{3}$$

A grid search minimising the cross-validation root mean square prediction error on a random sample of fourteen months worth of trap data determined the values for nuisance parameters $m_j$ (number of nearest neighbours), $C$ (a scalar denoting the relative importance of time to geographical distance), and $p$ (the inverse distance weighting factor).

### 3 Changes in WEI

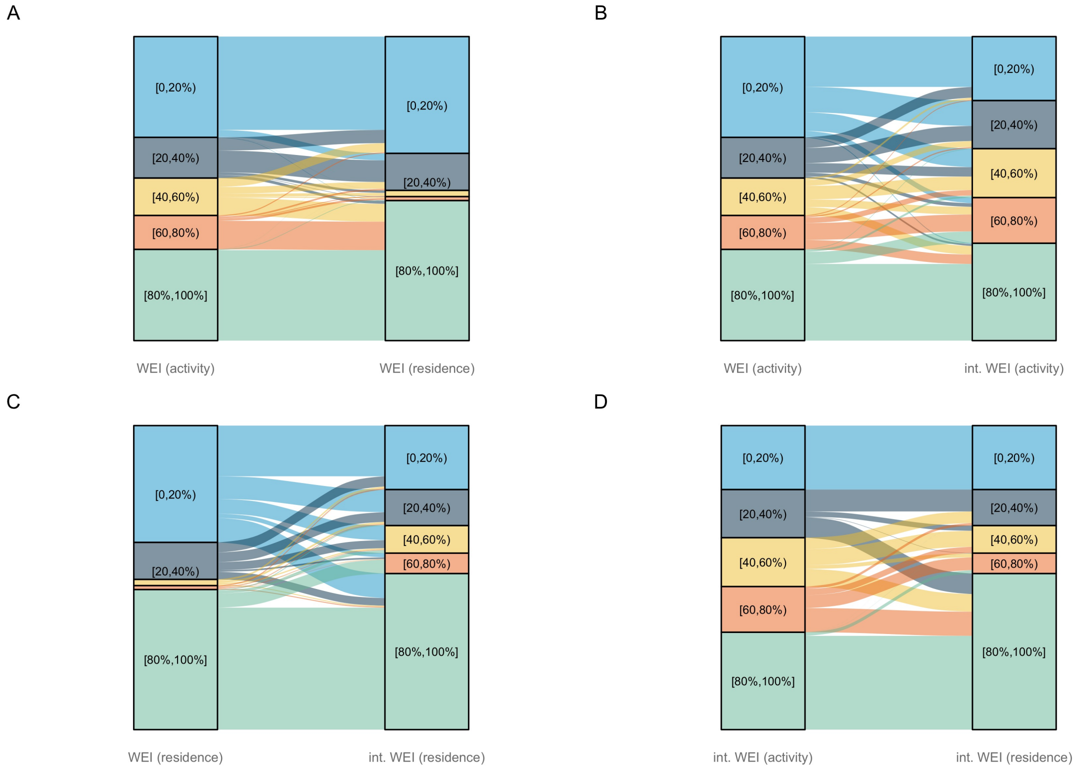


Figure S3: Changes in participant-level WEI values based on the method of estimation. A) The individual-level WEI values estimated in the original per-protocol analysis based on cluster-level *w*Mel prevalence during the month of enrollment, estimated either as a weighted average of prevalences in the clusters visited ("activity", left axis) or based on cluster-aggregated *w*Mel levels at the cluster of residence ("residence", right axis). B) The original per-protocol individual-level WEI (activity) values (left) are compared to the individual-level WEI (activity) values from the interpolated surface (right). C) The original per-protocol individual-level WEI (residence) values (left) are compared to the individual-level WEI (residence) values from the interpolated surface (right). C) A recreation of the plot in A, but using the interpolated WEI (activity) and interpolated WEI (residence) values.

### 4 Determining the "Compliers" for Complier-Restricted Analysis

The complier-restricted analysis restricted the analytic dataset to those who spent all reported time under the intervention assignment determined by their cluster of residence (n = 3,114). Time spent outside of the AWED study area was handled two ways. First, individuals who left the AWED trial area were excluded. Second, the analysis was broadened to include individuals whose time inside and outside of the AWED study area fell under their assigned intervention arm. For individuals in the intervention arm, this means including those who only spent time in the AWED intervention area and the quasi-experiment intervention area. For those in the untreated arm, this includes individuals who spent time anywhere outside of the AWED study area except for in the quasi-experiment intervention area.
